# A multicentre prospective observational study to investigate the prevalence and short-term impact of frailty, sarcopenia, anaemia and multiple long term health conditions in chronic limb threatening ischaemia (CLTI) - The FraiLTI (Frailty in Chronic Limb Threatening Ischaemia) Study

**DOI:** 10.1101/2024.11.11.24317086

**Authors:** Ashwin Sivaharan, Blaise Hickson, Tamer El-Sayed, Lauren Shelmerdine, Sai Wunnava, Emily James, Craig I Nesbitt, Alasdair O’Doherty, Miles D Witham, Sandip Nandhra

**Affiliations:** Northern Vascular Centre, Freeman Hospital, Freeman Rd, High Heaton, Newcastle upon Tyne, NE7 7DN; The Medical School, Newcastle University, Framlington Place, Newcastle upon Tyne, NE2 4HH; NIHR Leicester Biomedical Research Centre, Leicester, UK; Diabetes Research Centre, University of Leicester, Leicester, UK; Department of Sport and Exercise Sciences, Durham University, Green Ln, Durham, DH1 3LA; Department of Sport, Exercise and Rehabilitation, Northumbria University, Newcastle upon Tyne, NE1 8ST; AGE Research Group, Translational and Clinical Research Institute, Faculty of Medical Sciences, Newcastle University, Newcastle upon Tyne, UK; NIHR Newcastle Biomedical Research Centre, Newcastle upon Tyne Hospitals NHS Foundation Trust, Cumbria Northumberland Tyne and Wear NHS Foundation Trust and Faculty of Medical Sciences Newcastle University, Newcastle upon Tyne, UK; Population Health Sciences Institute, Faculty of Medical Sciences, Newcastle University, Baddiley-Clark Building, Newcastle upon Tyne, NE2 4AX; Newcastle upon Tyne Hospitals NHS Foundation Trust; South Tees Hospitals NHS Foundation Trust; University Hospitals Birmingham NHS Foundation Trust; NHS Lothian; University Hospitals of Leicester NHS Trust; Hull University Teaching Hospitals NHS Trust

**Author notes:** Authors share co-first authorship (Ashwin Sivaharan, Blaise Hickson). **Funding:** This work was funded by the National Institute for Health and Care Research (NIHR) Newcastle Biomedical Research Centre (reference: IS_BRC_1215_20001). The views expressed in this publication are those of the authors and do not necessarily reflect the views of the National Institute for Health and Care Research or the Department of Health and Social Care. <u>*Corresponding Author:*</u> Mr Ashwin Sivaharan, Northern Vascular Centre, Freeman Hospital, Freeman Rd, High Heaton, Newcastle upon Tyne, NE7 7DN. <u>Category:</u>* Observational study. <u>No conflicts of interest to declare</u>.

**Keywords:** Chronic limb threatening ischaemia (CLTI), frailty, sarcopenia, multiple long-term health conditions, anaemia

## Abstract

**Background:** CLTI is a life and limb threatening condition that is associated with a gradual overall decline in health which remains under-explored. We explore the prevalence of correlated conditions (frailty, sarcopenia and anaemia) in patients with CLTI and assess their ability to perform activities of daily living (ADLs) alongside multiple long term health conditions (MLTC), polypharmacy and clinical outcomes.

**Methods:** FraiLTI was a UK multi-centre, prospective, observational study, evaluating the prevalence of frailty (Rockwood Clinical Frailty score of ≥5), sarcopenia based on low handgrip strength (<27kg for men and <16kg for women). and anaemia in patients admitted to hospital with chronic limb threatening ischaemia (CLTI). Other outcomes included prevalence of polypharmacy (≥5 medications) and MLTC (≥2 long-term health conditions). Recruitment took place from October 2021 to October 2022 in six UK centres supported by the Vascular and Endovascular Research Network and funded by the National Institute for Health and Care Research (NIHR) Biomedical Research Centre (BRC) North-East.

**Results:** 84 patients were included in the study (54 [64.3%] men), with a mean (SD) age of 71.5 (11) years. 43/83 (51.8%) patients were living with frailty. 27/81 (33.3%) patients had probable sarcopenia 43/80 (53.8%) patients had anaemia of which 35 (92.1%) had normocytic anaemia. There was no difference in 90-day amputation-free survival for those living with or without frailty (97.6% vs 94.8%, p=0.78), sarcopenia (100% vs 94.4%, p=0.58), or anaemia (95.1% vs 97.3% vs p=0.45). MLTC were present in 57 (67.9%) patients and 67 (79.8%) patients met the definition of polypharmacy.

**Conclusion:** Frailty, sarcopenia and anaemia are highly prevalent in those with CLTI but none of these factors were associated with amputation free survival in this study. This has highlighted that the management of CLTI patients extends beyond revascularisation alone.

**Article Highlights:** *Type of research:* Multi-centre, prospective, observational study.

*Key Findings:* The prevalence of frailty (51.8%), anaemia (53.8%), and sarcopenia (33.3%) is high in people with CLTI. Most patients with CLTI had multiple long-term conditions (MLTC) (67.9%) or polypharmacy (79.8%).

*Take home Message:* This study shows that frailty, sarcopenia and anaemia are highly prevalent in those with CLTI. The management of CLTI patients extends beyond revascularisation alone.

**Table of contents Summary:** This study shows that frailty, sarcopenia and anaemia are highly prevalent in those with CLTI. The management of CLTI patients extends beyond revascularisation alone.

## Introduction

Chronic Limb Threatening Ischaemia (CLTI) has been highlighted as a research priority by both patients and clinicians, including in a recent James Lind Alliance (JLA) priority-setting partnership.^1^ CLTI is associated with a 3-year mortality rate of up to 40%, an increased risk of limb loss,^2^ and has a negative impact on patients’ quality of life.^3^ Patients with CLTI often have many associated conditions that might worsen outcomes.

Frailty is defined as an age-related multisystem impairment in homeostasis that leaves patients vulnerable to stressors such as illness, trauma, or surgery^4^. Frailty is related to nutritional deficiencies, the presence of multiple chronic conditions, hormonal changes and increased systemic inflammation.^5,6^ Frailty has been associated with worse outcomes in a wide range of surgical disciplines^7,8^, including vascular surgery.^9,10^ Sarcopenia is a distinct, but related concept, that describes age-related loss of muscle strength, muscle mass and muscle function.^11^ Sarcopenia forms an important component of the physical frailty syndrome ^12^ and has been associated with worse survival following endovascular surgery^13^ and surgical revascularisation.^14^

Multiple long-term conditions (MLTC; sometimes referred to as multimorbidity) may also be a contributing factor to outcomes in this population, both with respect to cardiovascular and non-cardiovascular conditions and has been associated with poorer outcomes in multiple surgical subspecialities.^15^ There are also retrospective data that suggest that anaemia, a potential marker of overall health state, is associated with worse outcomes in lower limb vascular bypass patients.^16^

Currently there exists no prospective data that describes the prevalence of these associated conditions in the population with CLTI.

FraiLTI (Frailty in chronic Limb Threatening Ischaemia) was a UK multi-centre prospective observational study aimed at evaluating the prevalence and impact of frailty (and associated conditions) on outcomes for patients presenting to hospital with CLTI.

## Methods

### Study design & setting

This was a multicentre, prospective observational study was conducted from October 2021 to October 2022, led by a Newcastle University based Principal Investigator team and supported by the Vascular and Endovascular Research Network (VERN). Recruitment was undertaken in 6 UK Vascular Centres, including, Newcastle upon Tyne Hospitals NHS Foundation Trust, South Tees Hospitals NHS Foundation Trust, University Hospitals Birmingham NHS Foundation Trust, University Hospitals of Leicester NHS Trust, Hull University Teaching Hospitals NHS Trust and NHS Lothian. The study was funded by the National Institute of Health Research Biomedical Research Centre (NIHR-BRC) and the Newcastle Hospitals Charity with full ethical approval (21/PR/0750 REC by the London - Hampstead NHS Research Ethics Committee). The study is registered on the ISRCTN registry (ISRCTN18644880) and the protocol has been published in full.^17^ The manuscript has been prepared as per the STROBE (Strengthening the reporting of observational studies in epidemiology) guidelines for the reporting of observational studies.^18^

### Participants

All patients admitted to UK vascular centres, with CLTI as per consensus definition (the presence of peripheral artery disease (PAD) in combination with rest pain, gangrene, or a lower limb ulceration >2 weeks duration)^19^ were invited to participate in the study across six sites. Patients were excluded if they were younger than 18 years, pregnant, or unable to consent to participation.

### Outcomes

The main outcomes assessed included frailty (see definitions), sarcopenia and anaemia. Other outcomes included walk speed, self-reported low physical activity, multiple long-term conditions and polypharmacy. Key clinical outcomes (measured at 90 days) included observed clinical care pathway (revascularisation/amputation etc.), mortality, major adverse limb events (MALEs), major adverse cardiovascular events (MACE), readmissions, reinterventions and other complications (including respiratory, urinary, and wound-related). Interaction between the main outcomes and the clinical outcomes were then analysed (see statistical analysis section).

#### Definitions

Frailty was assessed using the Rockwood CFS. A Rockwood CFS of ≥5 was considered frail (a score of 5 denotes mild frailty).^20^ Sarcopenia was assessed using grip strength, measured using a hand dynamometer (Jamar Smart Digital Hand Dynamometer). Participants were seated with the elbow flexed at ninety degrees and the wrist positioned with the thumb facing upwards. Six readings were taken in total (3 from each hand), and the highest reading was used for analysis. Grip strength of <27kg for men and <16kg for women was indicative of probable sarcopenia.^11^

Anaemia was defined as a haemoglobin of <130g/L in men and <120g/L in women as per the World Health Organisation (WHO) definition,^21^ based on the most recent pre-operative blood tests.

A low walk speed was diagnosed if the speed was <0.8m/s was noted on a 4m walk test. If the patient was unable to complete the test, this was considered diagnostic of low walk speed.

Low physical activity was measured using four activity questions used in the English Longitudinal Study of Ageing.^22^ The methodology used to diagnose low physical activity based on these questions has previously been published.^23^

MLTC was defined as ≥2 long-term health conditions (including ischaemic heart disease, previous stroke/ TIA, heart failure, atrial fibrillation, hypothyroidism, dementia, anxiety and depression, Parkinson’s disease, chronic obstructive pulmonary disease, asthma, osteoarthritis and any other inflammatory arthropathies).

Polypharmacy was defined as ≥5 medications, with excessive polypharmacy defined as ≥10 medications.^24^

Major adverse limb events (MALE) were defined as above ankle amputation of the index limb or major vascular reintervention.^25^ MACE was defined as a composite of acute myocardial infarction, stoke or cardiovascular death.^26^ Any lower limb reintervention or readmission for any cause within 90 days was recorded.

#### Data collection

Anonymised data included patient demographics, previous treatments, and hospital admissions from the past six months. The research teams gathered this information prospectively using both electronic and paper-based hospital records. The data was then entered into a custom-built, secure, electronic database, hosted on the Newcastle Joint Research Office’s Research Electronic Data Capture (REDCap) platform.

The following conditions were documented based on the patients’ medical histories: ischaemic heart disease or previous myocardial infarction, atrial fibrillation, hypertension, cerebrovascular diseases (such as ischaemic stroke, haemorrhagic stroke, or transient ischaemic attack), end-stage renal failure requiring dialysis, and chronic obstructive pulmonary disease. Pre-operative medications were noted from pre-assessment clinic records.

Routine clinical laboratory results (including haemoglobin [g/dl], white cell count [10^9^/L], albumin [g/L], creatinine [µmol/L], eGFR [mL/min/1.73 m²], C-reactive protein [mg/L], and HbA1c [mmol/mol]) were also collected, along with height and weight measurements.

#### Statistical methods

Statistical analysis was performed using R version 4.5.0 (R Foundation for Statistical Computing, Vienna, Austria). Descriptive proportional data is presented as number (percentage, 95% confidence interval (CI), calculated using the exact binomial test). Continuous data is presented as mean (SD), and hypothesis testing was performed with t-tests or Mann Whitney u-tests as appropriate based on normality testing with the Shapiro-Wilk test.

Proportional comparisons were performed with Fisher’s exact tests. A p-value <0.05 was considered statistically significant for single comparisons. Log rank tests are reported to compare the overall amputation-free survival (AFS).

## Results

Eighty-four participants were recruited between October 2021 to October 2022. 54 (64%) were men and the average BMI was 27.4 (6.9). 20 (24%) patients were current smokers, 47 (57%) were ex-smokers, and 16 (19%) were never-smokers. Demographics, co-morbidities and social factors are summarised in Table 1.

**Table 1:**
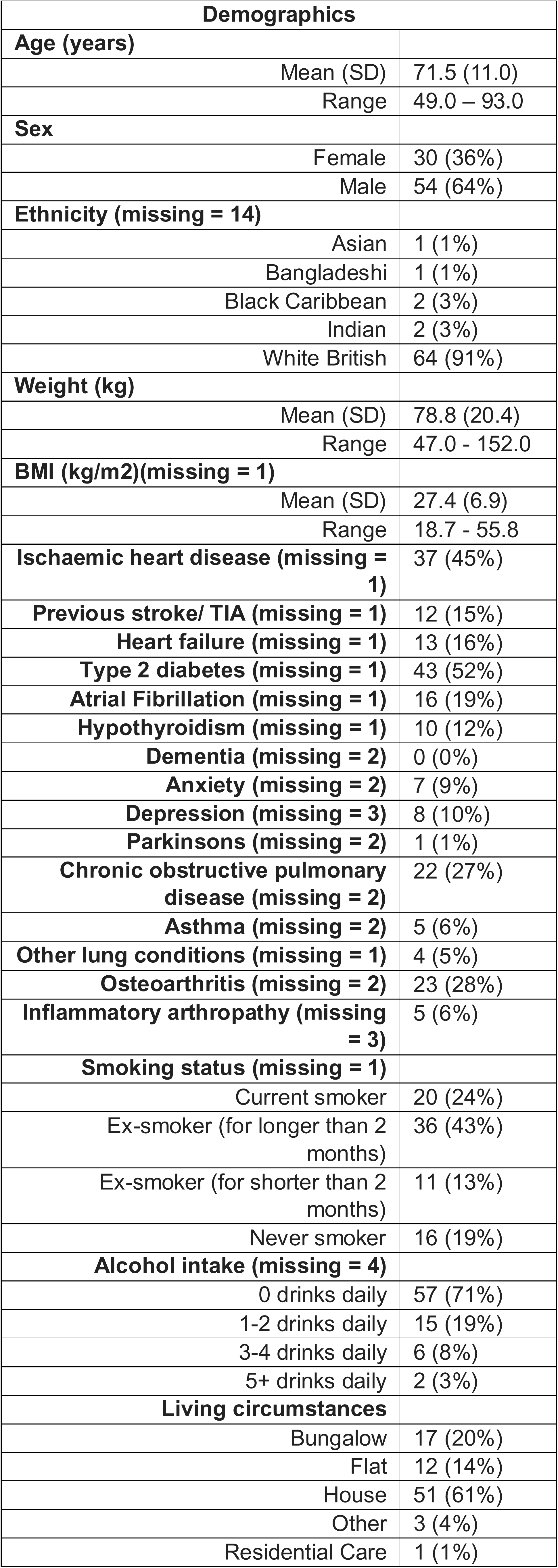
Table showing baseline demographics, co-morbidities and social history.

### Main outcomes (see table 2)

Fifty-two percent (43/83 participants) were living with frailty, with median (IQR) Rockwood CFS of 5(2). Patients who were living with frailty were more likely to be sarcopenic by grip strength (19/43 (46%) vs 8/40 (20%), p=0.018). These results are summarised in Table 3.

**Table 2:**
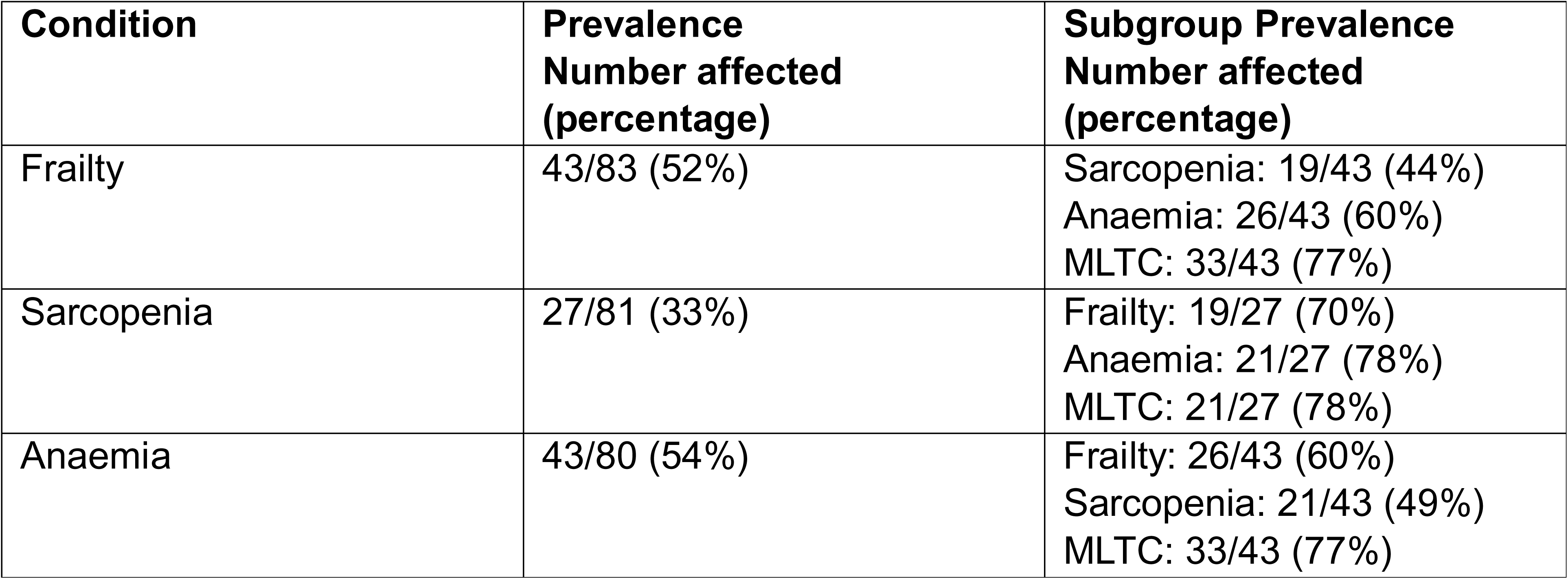
Table showing prevalence of main outcomes and the interaction between conditions.

| **Condition** | **Prevalence**<br>**Number affected**<br>**(percentage)** | **Subgroup Prevalence**<br>**Number affected**<br>**(percentage)** |
| --- | --- | --- |
| Frailty | 43/83 (52%) | Sarcopenia: 19/43 (44%)<br>Anaemia: 26/43 (60%)<br>MLTC: 33/43 (77%) |
| Sarcopenia | 27/81 (33%) | Frailty: 19/27 (70%)<br>Anaemia: 21/27 (78%)<br>MLTC: 21/27 (78%) |
| Anaemia | 43/80 (54%) | Frailty: 26/43 (60%)<br>Sarcopenia: 21/43 (49%)<br>MLTC: 33/43 (77%) |

**Table 3:** Table summarising the relationships between frailty and clinical factors. All numbers are reported to the nearest whole number (and percentage). Missing data values are included in square brackets. *Composite including MACE, MALE, respiratory, urinary or wound-related complications.

| **Clinical Frailty scale (missing = 1)** | **Not Frail**<br>**(N=40)** | **Frail**<br>**(N=43)** | **p value** |
| --- | --- | --- | --- |
| Sarcopenic (grip strength) | 8 (20%) | 19<br>(46%)<br>[2] | **0.018\*** |
| MLTC | 24 (60%) | 33<br>(77%) | 0.16 |
| Anaemia | 17 (44%)<br>[2] | 26<br>(63%)<br>[2] | 0.12 |
| Walk speed test failure | 10 (25%) | 21<br>(49%) | **0.040\*** |
| Low Activity | 17 (43%) | 39<br>(91%) | **<0.001\*\*\*** |
| Polypharmacy | 31 (78%) | 36<br>(84%) | 0.58 |
| **Revascularisation** |  |  |  |
| Revascularisation performed | 33 (85%)<br>[1] | 31<br>(74%)<br>[1] | 0.28 |
| Type of revascularisation | [7] | [13] | 0.20 |
| Endovascular | 16 (49%) | 20<br>(67%) |  |
| Open or Hybrid | 17 (52%) | 10<br>(33%) |  |
| **Complications** |  |  |  |
| MACE | 0 (0%)<br>[1] | 3 (7%)<br>[1] | 0.24 |
| MALE | 3 (8%)<br>[1] | 3 (7%)<br>[1] | 1.00 |
| All complications* | 4 (10%)<br>[1] | 9 (21%)<br>[1] | 0.23 |
| **Readmissions and reinterventions** |  |  |  |
| Number of readmission | [4] | [5] | 0.14 |
| 0 | 24 (67%) | 21<br>(55%) |  |
| 1 | 9 (25%) | 10<br>(26%) |  |
| 2 | 1 (3%) | 5 (13%) |  |
| 3 | 2 (6%) | 0 (0%) |  |
| 4 | 0 (0%) | 2 (5%)<br>[1] |  |
| Reintervention (Open or hybrid) | 3 (8%) [1] | 1 (2%)<br>[1] | 0.35 |
| Reintervention (Endovascular) | 1 (3%) [1] | 5 (12%)<br>[1] | 0.20 |
| Reintervention (Wound debridement) | 1 (3%) [1] | 1 (2%)<br>[1] | 1.00 |
| Reintervention (Foot debridement) | 4 (10%) [1] | 2 (5%)<br>[1] | 0.42 |

A third (33%) of participants (27/81) had sarcopenia by grip strength. Patients who were considered sarcopenic by grip strength were more likely to be anaemic (21/27 (79%) vs 21/54 (41%), p=0.004). These results are summarised in Supplementary Table 1.

Overall, 54% (43/80) were anaemic, with the majority being normocytic (35/38, 92%). Supplementary Table 2 summarises these findings.

For patients who were considered frail, 19/43 (46%) were sarcopenic by grip strength and 26/43 (63%) were anaemic.

#### Survival analysis

90-day AFS in the whole cohort was 96%. There was no significant difference in 90-day AFS for those living with frailty (98% vs 95%, p=0.78), sarcopenia (100% vs 94%, p=0.58), or anaemia (95% vs 97%, p=0.45) or MLTC (96 vs 96%, p=0.67).

#### Other outcomes

Low walk speed (<0.8m/s) or failure of the 4m walk speed test was observed in 30/82 patients (37%). The self-reported physical activity index suggested that 56 patients (68%) were classified as having low activity. MLTC was seen in 57 (68%) patients and 67 (80%) patients met the definition of polypharmacy.

Patients living with frailty were more likely to fail the walk speed test (21/43 (49%) vs 10/40 (25%), p=0.040) and self-report low activity (39/43 (91%) vs 17/40 (43%), p<0.001). Anaemic patients had a greater reduction in activity levels (35/43 (81%) vs 19/37 (51%), p=0.008) and were more likely to fail the walk speed test (23/43 (54%) vs 8/37 (22%), p=0.005).

Pearson correlation coefficient was computed to determine the relationship between haemoglobin and grip strength. The results suggest a positive relationship that was statistically significant (r(76) = 0.35, p=0.0017). Figure 2 shows a scatterplot demonstrating this relationship.

**Figure 1:**
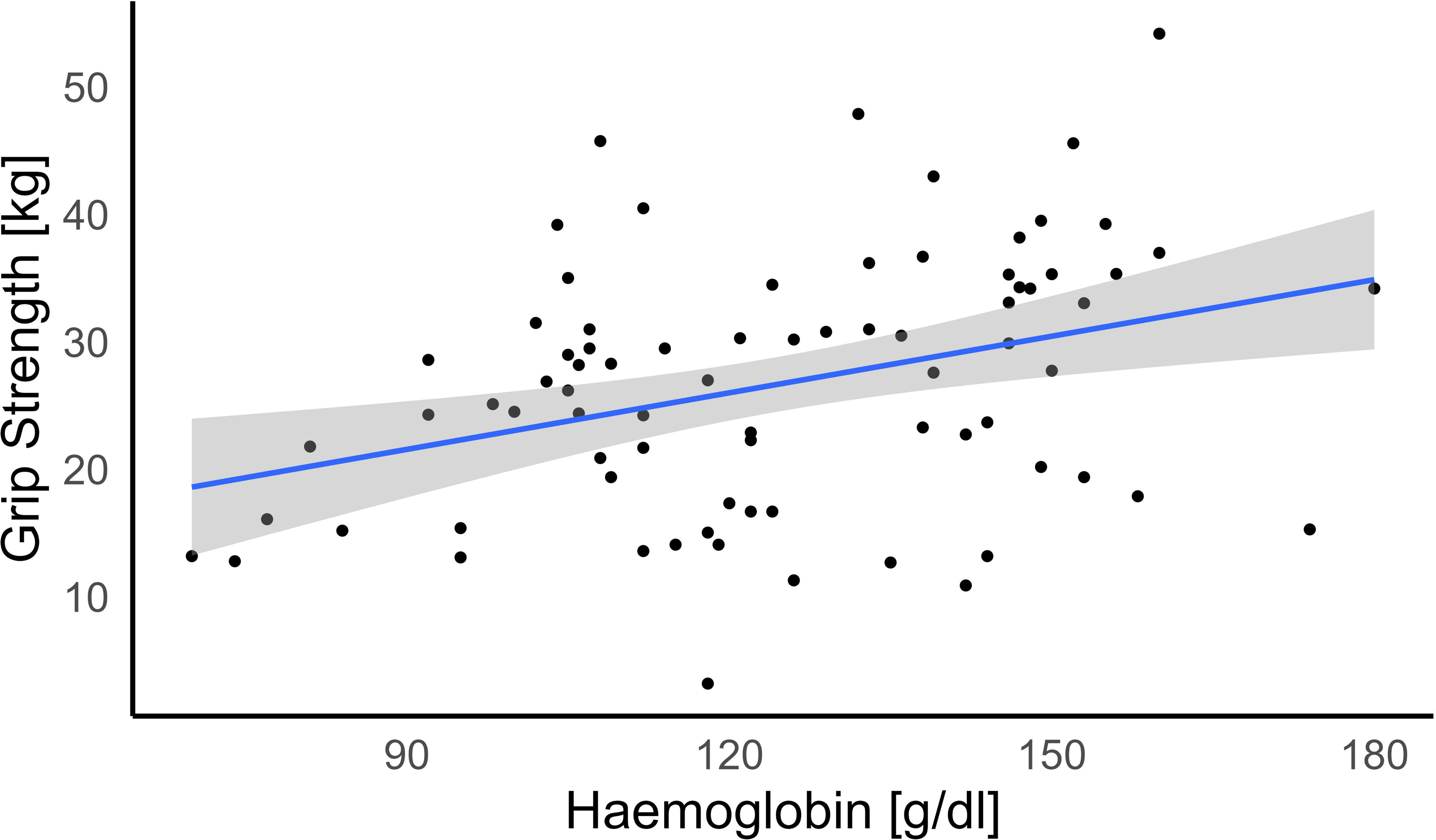
Scatterplot showing relationship between haemoglobin and grip strength.

**Figure 2:**
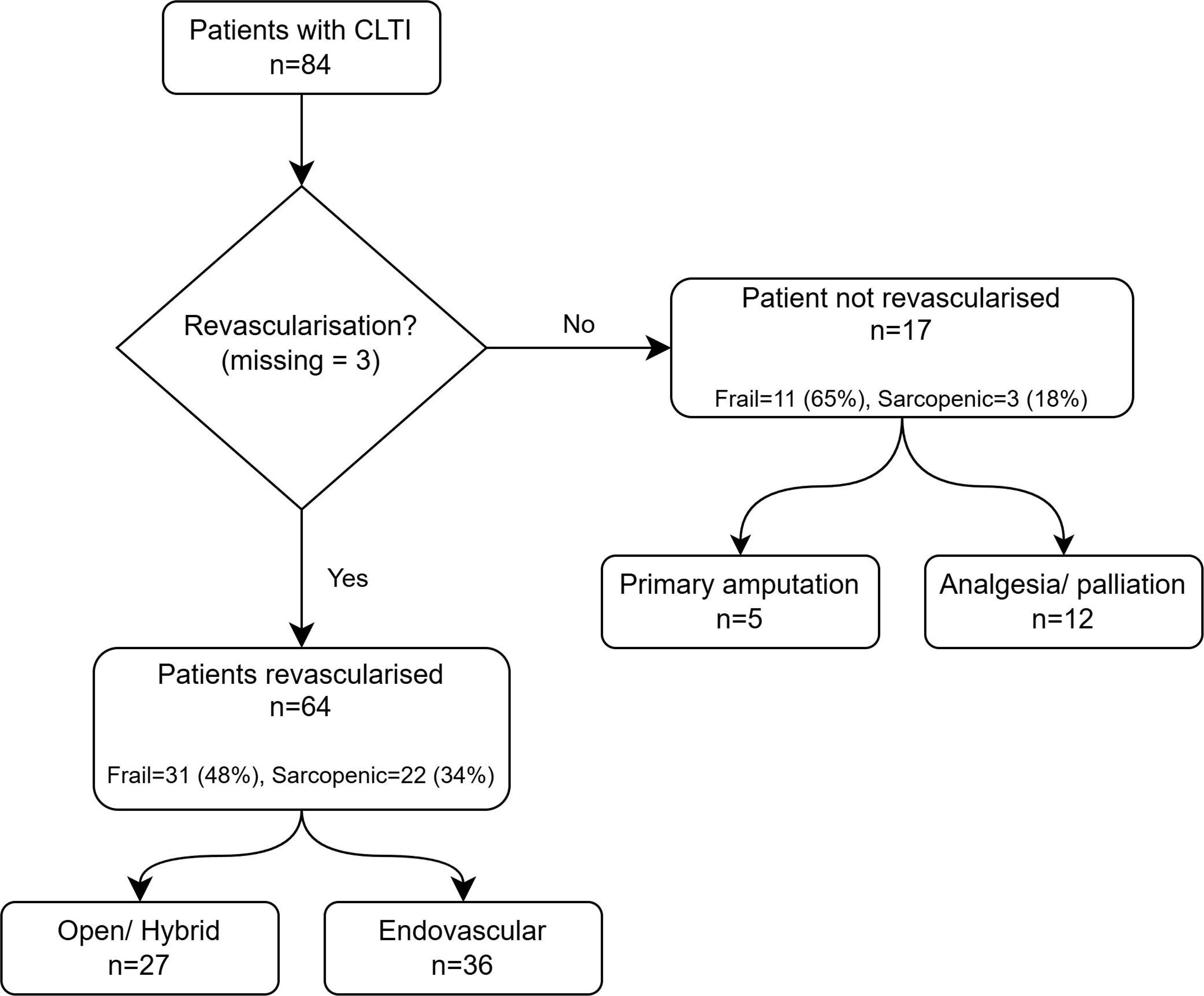
Flowchart showing how many patients underwent intervention for CLTI. There were 3 missing data points for revascularisation.

#### Clinical outcomes

Most patients (64/81; 79.0%) underwent revascularisation, 36 (57.1%) were endovascular and 27 (42.9%) open or hybrid. Seventeen (21%) patients did not undergo revascularisation; 5 (29.4%) underwent primary amputation, and 12 (70.6%) underwent a conservative or palliative approach. Figure 2 shows a flowchart summarising this. Those with anaemia were less likely to undergo any open/hybrid reinterventions within 90 days (0 (0.0%) vs 4 (10.8%), p=0.046), but more likely to undergo minor foot debridement (6 (14.6%) vs 0 (0.0%), p=0.027).

During the study period, there were 3 (4%) MACEs, 2 (3%) respiratory complications, 5 (6%) wound complications and 6 (7%) MALEs. 29 (35%) patients in the study had at least one readmission while in the study.

Sixteen patients had reinterventions within 90 days of index surgery; 4 (5%) open/ hybrid interventions, 6 (7%) endovascular interventions, 2 (2%), wound-related procedures and 6 (7%) foot debridements/ minor amputations.

### Relationships between frailty, sarcopenia, anaemia and clinical factors

There were no significant correlations found between the presence of frailty and the complication rate, readmission rates or the number and type of reinterventions. These results are summarised in Table 3.

Supplementary tables 3 and 4 summarise the relationships between walk speed test failure and self-reported low activity, against other characteristics.

## Discussion

In the first UK prospective observational study of its kind, we have shown that a high prevalence of frailty, sarcopenia, anaemia, MLTC and polypharmacy exists in the CLTI and PAD population. We have also shown that there were positive associations between frailty and sarcopenia and between anaemia and sarcopenia, among CLTI patients.

The high prevalence of frailty in the population is striking but unsurprising. Indeed, this is in keeping with the literature in the UK; a recent retrospective study has shown that 98/190 (52%) patients were living with frailty by the Rockwood CFS.^27^ A meta-analysis of global literature has suggested that 49% of those with PAD are frail, although this study was limited due to significant study heterogeneity.^28^ In comparison, the prevalence of frailty in English adults aged >50 years old, was 8.1% [95% CI 7.3-8.8].^29^

Our findings on the prevalence of sarcopenia also matches up reasonably with the literature, with previous retrospective data suggesting prevalence of around 40% (37.8%^30^ to 40.8%^31^). It is worth noting that most of these studies only include patients who undergo intervention, in contrast to our study, which includes those managed conservatively. Similarly, this study supports previously published findings of the prevalence of anaemia in the CLTI population of around 50%.^32^

Our study has identified an association between frailty and sarcopenia. The claudication symptoms that often precede CLTI can limit activity levels^33^ and by the time patients present with rest pain or tissue loss, this insidious reduction in physical activity may lead to loss of independence, functional muscle loss and sarcopenia.^34,35^ This poses the question of whether early revascularisation in those with claudication is warranted; there is reasonable data to support the fact that early surgery in this population leads to a higher risk of MLLA.^36^ This, therefore, leaves us with more conservative measures; aggressive lifestyle modification and supervised exercise programmes (usually walking and treadmill based regimes) reduce the progression and severity of claudication symptoms and improve quality of life.^37^ Unfortunately, these programmes are rarely available in the National Health Service.^38^ There is also a role for resistance training exercise regimes, as this type of exercise programme has been shown to be beneficial in those with sarcopenia.^39^ This could be incorporated into both pre-operative and post-operative rehabilitation strategies.

The description of anaemia and its relationship with sarcopenia by grip strength is a new finding in the UK CLTI population, however, it has been previously described in other non-vascular surgery populations, particularly in East Asia.^40–42^ Both anaemia and sarcopenia have been suggested to contribute to poor outcomes after vascular surgery.^14,16^ There is probably an element of mixed anaemia in this patient cohort, but there are certainly unanswered questions about the true mechanism of anaemia in this population.

Frailty and sarcopenia are both well understood to be part of aging and the concept of “inflammaging” has been suggested as a mechanism for this. The imbalance of pro- and anti-inflammatory processes are thought to drive this and result in inflammation mediated loss of muscle mass, and inhibition of muscle regeneration.^43^ There is potential that the disease of CLTI itself, and the associated local lower limb inflammation could be further driving this inflammatory state. It could be hypothesised that this could be the case with earlier stage PAD, where the onset of claudication triggers a systemic inflammatory response. Cellular senescence and mitochondrial dysfunction have also been postulated to play a role in the pathophysiology of sarcopenia.^44^

With such high rates of frailty, sarcopenia and anaemia in the vascular surgery population, it is incumbent that services take steps to manage, and where possible, treat these conditions. The main challenge to achieve this comes down to the short timeframes required for CLTI intervention (less than 14 days as an out-patient and less than 5 days as an in-patient).^45^ Complex geriatric assessment (CGA) is an increasingly used service in vascular surgery units and can be implemented via an inpatient or outpatient basis.^46–48^ Indeed, the increasing use of “hot clinics” and outpatient pathways may alleviate the risk of the deconditioning that often occurs with a prolonged hospital admission.^9^

### Limitations

Despite this being a UK-first, there remain some limitations of this study. Primarily, a lack of long-term follow-up limited our ability to draw conclusions regarding frailty, sarcopenia and their effect on long-term outcomes, such as amputation free survival.

In this study, there was no detectable relationship between frailty, sarcopenia or anaemia against survival or other major clinical outcomes such as amputation, readmission or death. This may represent that these factors do not affect mid-term mortality and amputation rates, however, this is more likely to be because our study cohort of 84 patients provides insufficient power to detect statistically significant rates of complications between groups. This issue was compounded by the low rates of complications and death in the study, a fact that probably reflects the short follow-up time and the often-reported problem of study patients not necessarily representing the real-world population.^49^

In addition, some of the planned analyses were limited due to missing data. The Fried Frailty index was a significant outcome which was limited in this way, mostly driven by a lack of data pertaining to unintentional weight loss. Skeletal muscle area at L3 was also affected, and this may represent technical challenges in the measurement of this variable, which could be alleviated in future studies by improved training, or perhaps centralised image analysis. This prevented some of the analyses that were initially outlined in the protocol.^17^ and impacted our ability to assess muscle quantity.

Future work to address the issues raised by this study should target the multiple facets of frailty and sarcopenia. Investigation of the mechanisms of sarcopenia and its relationship with anaemia, may lead to novel findings that could provide potential targets to slow down their progression. This approach may also unveil biological processes that are unique to the pathophysiology of CLTI.

Nutritional assessment and post-operative exercise and rehabilitation programmes may help alleviate the negative consequences of frailty and sarcopenia. Correcting anaemia may also be a potential avenue, and our unit is exploring further research on this topic.

## Conclusion

This prospective multi-centre study highlights that there is a high prevalence of frailty, sarcopenia and anaemia in a population with CLTI. The positive association between sarcopenia and anaemia is novel in this population, and merits rigorous evaluation in future research. Our results suggest that a focus should be placed on managing these conditions alongside CLTI in a multidisciplinary fashion, and there is scope for further larger-scale prospective studies on these conditions and their impact on outcomes and quality of life after arterial intervention.

## Data Availability

All data produced in the present study are available upon reasonable request to the authors

---

Supplementary Table 1: Table summarising the relationships between sarcopenia and clinical factors. All numbers are reported to the nearest whole number (and percentage). Missing data values are included in square brackets. *Composite including MACE, MALE, respiratory, urinary or wound-related complications.

Supplementary Table 2: Table summarising the relationships between anaemia and clinical factors. All numbers are reported to the nearest whole number (and percentage). Missing data values are included in square brackets. *Composite including MACE, MALE, respiratory, urinary or wound-related complications.

Supplementary Table 3: Table summarising the relationships between walk speed and clinical factors. All numbers are reported to the nearest whole number (and percentage). Missing data values are included in square brackets. *Composite including MACE, MALE, respiratory, urinary or wound-related complications.

Supplementary Table 4: Table summarising the relationships between self-reported low activity and clinical factors. All numbers are reported to the nearest whole number (and percentage). Missing data values are included in square brackets. *Composite including MACE, MALE, respiratory, urinary or wound-related complications.

## Acknowledgements

The authors of this study would like to acknowledge the involvement of the vascular research team (Samantha Mallon, Noala Parr, Esther Olabiyi and Tony Robinson), who provided invaluable assistance in the running of the study, and the clinical surgical teams in all recruitment sites, who supported recruitment to the study. In addition, we would like to recognise the contribution of the Vascular and Endovascular Research Network (VERN) for their support in running the study.

MDW acknowledges support from the NIHR Newcastle Biomedical Research Centre. SN acknowledges support from the NIHR Academy and the Newcastle Health Innovation/Research Partnership.

