## Supplementary table 1 for "A multicentre prospective observational study to investigate the prevalence and short-term impact of frailty, sarcopenia, anaemia and multiple long term health conditions in chronic limb threatening ischaemia (CLTI) - The FraiLTI (Frailty in Chronic Limb Threatening Ischaemia) Study"

| **Sarcopenia – grip strength**  **(missing = 3)** | **Not sarcopenic (N=54)** | **Sarcopenic (N=27)** | **p value** |
| --- | --- | --- | --- |
| **MLTC** | 34 (63%) | 21 (78%) | 0.21 |
| **Anaemia** | 21 (41%)  *[3]* | 21 (78%) | **0.004**** |
| **Walk speed test failure** | 16 (30%) | 13 (48%) | 0.14 |
| **Low Activity** | 32 (59%) | 22 (82%) | 0.051 |
| **Polypharmacy** | 43 (80%) | 23 (85%) | 0.76 |
| **Revascularisation** | | | |
| **Revascularisation performed** | 41 (76%) | 22 (88%)  *[2]* | 0.25 |
| **Type of revascularisation** | *[13]* | *[6]* | 0.11 |
| Endovascular | 20 (49%) | 15 (71%) |  |
| Open or Hybrid | 21 (51%) | 6 (29%) |  |
| **Complications** | | | |
| **MACE** | 2 (4%) | 1 (4%)  *[2]* | 1.00 |
| **MALE** | 4 (7%) | 2 (8%)  *[2]* | 1.00 |
| **All complications*** | 9 (17%) | 9 (17%)  *[2]* | 1.00 |
| **Readmissions and reinterventions** | | | |
| **Number of readmission** | *[4]* | *[3]* | 0.40 |
| 0 | 31 (62%) | 14 (58%) |  |
| 1 | 13 (26%) | 6 (25%) |  |
| 2 | 4 (8%) | 2 (8%) |  |
| 3 | 2 (4%) | 0 (0%) |  |
| 4 | 0 (0%) | 2 (8%) |  |
| **Reintervention (Open or hybrid) (missing = 2)** | 4 (7%) | 0 (0%)  *[2]* | 0.30 |
| **Reintervention (Endovascular) (missing = 2)** | 2 (4%) | 4 (16%)  *[2]* | 0.076 |
| **Reintervention (Wound debridement) (missing = 2)** | 1 (2%) | 1 (4%)  *[2]* | 0.54 |
| **Reintervention (Foot debridement) (missing = 2)** | 3 (6%) | 3 (12%)  *[2]* | 0.37 |
