## Supplementary table 2 for "A multicentre prospective observational study to investigate the prevalence and short-term impact of frailty, sarcopenia, anaemia and multiple long term health conditions in chronic limb threatening ischaemia (CLTI) - The FraiLTI (Frailty in Chronic Limb Threatening Ischaemia) Study"

| **Anaemia**  **(missing = 4)** | **Not anaemic (N=37)** | **Anaemic (N=43)** | **p value** |
| --- | --- | --- | --- |
| **Polypharmacy** | 28 (76%) | 36 (84%) | 0.41 |
| **MLTC** | 22 (60%) | 33 (77%) | 0.15 |
| **Walk speed test failure** | 8 (22%) | 23 (54%) | **0.005**** |
| **Low Activity** | 19 (51%) | 35 (81%) | **0.008**** |
| **Revascularisation** | | | |
| **Revascularisation performed** | 31 (84%) | 31 (76%)  *[2]* | 0.41 |
| **Type of revascularisation** | *[6]* | *[13]* | **0.004**** |
| Endovascular | 12 (39%) | 23 (77%) |  |
| Open or Hybrid | 19 (61%) | 7 (23%) |  |
| **Complications (missing = 2)** | | | |
| **MACE** | 2 (6%) | 1 (2%)  *[2]* | 0.60 |
| **MALE** | 2 (6%) | 4 (10%)  *[2]* | 0.68 |
| **All complications*** | 6 (16.2%) | 6 (14.0%)  *[2]* | 1.00 |
| **Readmissions and reinterventions** | | | |
| **Number of readmission** | *[2]* | *[7]* | 0.73 |
| 0 | 21 (60%) | 22 (61%) |  |
| 1 | 10 (29%) | 9 (25%) |  |
| 2 | 3 (9%) | 3 (8%) |  |
| 3 | 1 (2.9%) | 0 (0.0%) |  |
| 4 | 0 (0.0%) | 2 (5.6%) |  |
| **Reintervention (Open or hybrid) (missing = 2)** | 4 (10.8%) | 0 (0.0%)  *[2]* | **0.046*** |
| **Reintervention (Endovascular) (missing = 2)** | 2 (5%) | 4 (10%)  *[2]* | 0.68 |
| **Reintervention (Wound debridement) (missing = 2)** | 0 (0 %) | 1 (2%)  *[2]* | 1.000 |
| **Reintervention (Foot debridement) (missing = 2)** | 0 (0%) | 6 (15%)  *[2]* | **0.027*** |
