## Supplementary table 3 for "A multicentre prospective observational study to investigate the prevalence and short-term impact of frailty, sarcopenia, anaemia and multiple long term health conditions in chronic limb threatening ischaemia (CLTI) - The FraiLTI (Frailty in Chronic Limb Threatening Ischaemia) Study"

| **Walk speed test** | **Successful (N=52)** | **Unsuccessful (N=32)** | **p value** |
| --- | --- | --- | --- |
| **Polypharmacy** | 40 (77%) | 27 (84) | 0.58 |
| **MLTC** | 35 (67%) | 22 (69%) | 1.00 |
| **Anaemia** | 20 (41%)  *[3]* | 23 (74%)  *[1]* | **0.005**** |
| **Low Activity** | 29 (56%) | 27 (87%)  *[1]* | **0.004**** |
| **Revascularisation** | | | |
| **Revascularisation performed** | 44 (86%)  *[1]* | 20 (67%)  *[2]* | **0.049*** |
| **Type of revascularisation** | *[9]* | *[12]* | 0.43 |
| Endovascular | 23 (54%) | 13 (65%) |  |
| Open or Hybrid | 20 (47%) | 7 (35%) |  |
| **Complications** | | | |
| **MACE** | 3 (6%)  *[1]* | 0 (0%)  *[2]* | 0.29 |
| **MALE** | 3 (6%)  *[1]* | 3 (10%)  *[2]* | 0.67 |
| **All complications*** | 8 (15%)  *[1]* | 5 (16%)  *[2]* | 1.00 |
| **Readmissions and reinterventions** | | | |
| **Number of readmission** | *[4]* | *[6]* | 0.62 |
| 0 | 28 (58%) | 17 (65%) |  |
| 1 | 14 (29%) | 5 (19%) |  |
| 2 | 3 (6%) | 3 (12%) |  |
| 3 | 2 (4%) | 0 (0%) |  |
| 4 | 1 (2%) | 1 (4%) |  |
| **Reintervention (Open or hybrid)** | 3 (6%)  *[1]* | 1 (3%)  *[2]* | 1.00 |
| **Reintervention (Endovascular)** | 5 (10%)  *[1]* | 1 (3%)  *[2]* | 0.41 |
| **Reintervention (Wound debridement)** | 1 (2%)  *[1]* | 1 (3%)  *[2]* | 1.00 |
| **Reintervention (Foot debridement)** | 3 (6%)  *[1]* | 3 (10%)  *[2]* | 0.67 |
