## Supplementary table 4 for "A multicentre prospective observational study to investigate the prevalence and short-term impact of frailty, sarcopenia, anaemia and multiple long term health conditions in chronic limb threatening ischaemia (CLTI) - The FraiLTI (Frailty in Chronic Limb Threatening Ischaemia) Study"

| **Self-reported low activity (missing = 1)** | **False (N=27)** | **True (N=56)** | **p value** |
| --- | --- | --- | --- |
| **Polypharmacy** | 20 (74%) | 47 (84%) | 0.37 |
| **MLTC** | 17 (63%) | 40 (71%) | 0.46 |
| **Anaemia** | 8 (31%)  *[1]* | 35 (65%)  *[1]* | **0.008**** |
| **Revascularisation** | | | |
| **Revascularisation performed** | 24 (89%) | 40 (74%)  *[2]* | 0.16 |
| **Type of revascularisation** | *[3]* | *[17]* | 0.80 |
| Endovascular | 13 (54%) | 23 (59%) |  |
| Open or Hybrid | 11 (46%) | 16 (41%) |  |
| **Complications** | | | |
| **MACE** | 1 (4%) | 2 (4%)  *[2]* | 1.00 |
| **MALE** | 3 (11%) | 3 (6%)  *[2]* | 0.40 |
| **All complications*** | 5 (19%) | 8 (14%)  *[2]* | 0.75 |
| **Readmissions and reinterventions** | | | |
| **Number of readmission** |  | *[9]* | 0.21 |
| 0 | 14 (52%) | 31 (66%) |  |
| 1 | 9 (33%) | 10 (21%) |  |
| 2 | 2 (7%) | 4 (0%) |  |
| 3 | 2 (7%) | 0 (0%) |  |
| 4 | 0 (0%) | 2 (4%) |  |
| **Reintervention (Open or hybrid)** | 4 (15%) | 0 (0%)  *[2]* | **0.011*** |
| **Reintervention (Endovascular)** | 0 (0%) | 6 (11%)  *[2]* | 0.17 |
| **Reintervention (Wound debridement)** | 1 (4%) | 1 (2%)  *[2]* | 1.00 |
| **Reintervention (Foot debridement)** | 3 (11%) | 3 (6%)  *[2]* | 0.40 |
